# Leveraging blood RNA expression to understand Parkinson’s disease heterogeneity and progression

**DOI:** 10.64898/2025.12.10.25342029

**Authors:** Ana-Luisa Gil-Martinez, Aine Fairbrother-Browne, Raquel Real, Guillermo Rocamora-Perez, Jonathan W. Brenton, Alejandro Martinez-Carrasco, Juan A. Botia, Mina Ryten, Huw R Morris

## Abstract

Parkinson’s disease (PD) exhibits substantial clinical and molecular heterogeneity, driven in part by mutations in *SNCA*, *GBA1* and *LRRK2* genes. Understanding how these genetic subtypes differ in progression and peripheral transcriptomic profiles may inform personalized prognostic and therapeutic strategies. In this study, we aim to compare disease progression trajectories and blood-based gene expression patterns among *SNCA*-positive (*SNCA+*), *GBA*-positive (*GBA+*) and *LRRK2*-positive (*LRRK2+*), and sporadic PD patients, and to identify shared and subtype-specific transcriptional signatures. We used well-characterized cohorts of *SNCA*+ (n=26), *GBA*+ (n=70), *LRRK2*+ (n=147), and sporadic (n=387) PD patient from the Parkinson’s Progression Markers Initiative (PPMI) cohort. We used data from samples collected 6 months after baseline visit for differentially expressed gene (DEGs) analysis, controlling for cell type proportions. Our results show that *SNCA*+ patients had the earliest onset and poorest survival, whereas *LRRK2*+ and sporadic PD progressed more slowly. *GBA*+ and *LRRK2*+ patient groups exhibited prominent immune-related activation, with overlapping upregulated DEGs between *LRRK2*+ and *GBA*+ and shared downregulation between *LRRK2*+ and *SNCA*+. No significant blood DEGs distinguished favourable and unfavourable outcomes in sporadic PD cases. In conclusion, mutation-specific clinical trajectories and systemic inflammatory signatures in *GBA*+ and *LRRK2*+ PD underscore the promise of blood-based biomarkers, warranting validation in larger and tissue-targeted studies.

## INTRODUCTION

Parkinson’s disease (PD) is a complex neurodegenerative disorder that exhibits heterogeneity in its clinical characteristics, age at onset, progression and treatment response^1^. Emerging evidence increasingly challenges the traditional, simplified notion of PD as a disease defined exclusively by parkinsonism (motor symptoms), the presence of Lewy bodies, and dopaminergic neuronal loss in the substantia nigra. Such observations underscore that each individual evolves a specific form of the syndrome, defying simple classification schemes based strictly on clinical presentation or neuropathology. Consequently, there is an urgent need to determine and understand PD subtypes with different clinical manifestation and progression to facilitate more specific and timely intervention.

This complexity arises from an interplay of clinical, pathological and genetic factors. Clinically, PD patients exhibit a diverse range of motor symptoms and non-motor symptoms that can vary between individuals^3^. For instance, some classifications distinguish patients with tremor dominant PD from other cases with postural instability and gait difficulty. Non-motor features such as sleep disturbances, neuropsychiatric symptoms, and cognitive impairment contribute to the broad spectrum of PD subtypes. Moreover, identification of PD at the early stages of the disease is a great clinical challenge to distinguish it from other movement disorders such as multiple system atrophy, progressive supranuclear palsy and corticobasal degeneration with a reported error rate in clinical diagnosis in 7-35% of cases^4^.

The clinical heterogeneity of PD is also mirrored in Genome-Wide Association Studies (GWAS), which have revealed a complex genetic landscape contributing to the diverse manifestations of this disorder. Several studies have identified over 100 loci and numerous genetic variants associated with PD, some of which have been linked to different clinical PD phenotypes^5,6^. Genetic predisposition ranges from common risk alleles of modest effect to highly penetrant mutations (e.g. *SNCA*, *GBA* and *LRRK2*) that drive distinct trajectories and neuropathological signatures. Recent GWAS for progression have highlighted the potential role of genetic variation in driving disease phenotype and progression^7,8^. These genotype-phenotype correlations underscore the necessity of distinguishing hereditary PD subgroups from sporadic cases, both to refine prognostic models and to tailor therapeutic interventions.

The exploration of clinical and genetic variations between individuals diagnosed with PD has led to the development of advanced data-driven analysis techniques. Interestingly, Kristen Severson and colleagues developed a statistical progression model, accounting for intra-individual and inter-individual variability and medication effects, that showed non-sequential and overlapping disease progression trajectories. They indicated that a static subtype classification might be ineffective at capturing the complex heterogeneity of PD progression ^9^. These observations are in line with a previous study focused on the determination of the frequency and stability of PD subtypes in de novo patients. They concluded that instability must be taken into consideration specifically when establishing correlations with the biomarkers and for long term prognostication^10^.

RNA-seq data analysis has significantly advanced our understanding of the molecular mechanisms involved in various diseases, enabling the identification of key genetic markers and potential therapeutic targets. Specifically, the use of transcriptomic data has emerged as a powerful tool for investigating the progression of PD. Here, we used peripheral-blood RNAseq data from the Parkinson’s Progression Markers Initiative (PPMI) to identify transcriptomic profiles related to specific genetically defined PD patient subgroups. In addition, we considered PPMI-reported clinical features, such as age at diagnosis, disease duration, dementia and mortality to determine transcriptomic profiles and associated molecular mechanisms related with the subsequent development of a severe trajectory. With this study, we move beyond traditional case-control studies to prioritize PD subtype-specific mechanisms. The integration of expression data with genetic and clinical data can build a more comprehensive map of disease mechanisms and identify novel candidate genes for functional validation.

## METHODS

### Participant selection and unfavourable outcome criteria

We used data from the Parkinson’s Progression Markers Initiative (PPMI). We accessed data using the Accelerating Medical Partnership in Parkinson’s disease database (AMP-PD; https://www.amp-pd.org/ in which individuals are distributed within study arms determined by the inclusion/exclusion criteria established in the study protocol of each cohort. We selected participants from the following study arms: Genetic Cohort and Parkinson’s Disease (PD).

We pre-selected participants from the complete PPMI cohort (n=4,287, including controls) through a stepwise exclusion process. First, we removed individuals who declined participation or failed screening (n=1,402). Next, we retained only those with a Parkinson’s disease (PD) diagnosis at their last visit (n=1,385). We then excluded cases diagnosed with dementia with Lewy bodies within one year of PD onset (1,384), as well as individuals with fewer than two visits or overall follow-up shorter than one year (n=1,133). Finally, we excluded PD whose diagnosis changed during follow-up (n=1,130), yielding a final sample of n=1,130 PD cases.

Unfavourable diagnosis was defined according to criteria adapted from Manuela Tan *et al.*^7^ and Raquel Real *et al.*^8^. We categorized PD participants as having unfavourable outcome if they met one or more of the following: (i) mortality, death after study enrolment (“WDRSN” == 3), (ii) motor progression, Hoehn & Yahr stage >= 3 at any point (including baseline); and, (iii) cognitive impairment, incident dementia at any time point (including baseline) or withdrawal due to dementia.

### Kaplan-Meier survival analysis and Cox Proportional Hazards Modelling

For the Kaplan-Meier survival analysis and Cox proportional hazards regression, we used the R packages survival (version 3.8)^11^ and survminer (version 0.4.9)^12^. We generated the Kaplan-Meier survival curve with the survfit function, specifying disease duration and outcome status variables. Curves were stratified by group (*SNCA*+, *GBA*+, *LRRK2*+ and sporadic PD), and differences between strata were assessed via the log-rank test. We rendered the survival plot using the ggsurvplot function, with 95% confidence bands, p-value annotation, and a risk table below the x-axis to display the number of individuals per group at risk at each time point. To quantify the effect of each PD genetic group on the hazard of unfavourable outcome, we adjusted for potential confounders based on clinical relevance including age at baseline and sex and, we fitted a multivariable Cox model using coxph function.

### Whole-blood RNA-seq data

Individuals within each PPMI study arm underwent separate sample collection schemes, which carried distinct inclusion and exclusion criteria. Detailed documentation about sample collection and methodology is available on the PPMI website (https://www.ppmi-info.org). Samples were collected at various time-points corresponding to the number of months that the visit occurred after the baseline visit. For this study, we based our analysis on data collected 6 months after the baseline visit (M6), when the sample size was sufficiently large and well balanced to ensure adequate statistical power across PD subtypes.

### Covariate correction pipeline

We assessed sample and sequencing covariates for collinearity using a custom script to reduce the number of covariates tested. This used the stats (v4.2.3) and Hmisc (v5.2-0) packages to generate Spearman correlations, Kruskal-Wallis and chi-square tests for the assessment of relationships between covariates. When assessing numeric covariates that were collinear, those with the lowest overall correlations to all other variables were retained. We used VariancePartition (v1.32.5) scoring and correlations between the remaining covariates and the top 10 principal components (PCs) to assess which covariates should be controlled for in the final analysis. We also identified outlier samples were using these dataset-dependent covariates. Covariate-corrected gene counts, corrected using the limma46 (v3.58.1) removeBatchEffect function, were assessed. Samples that had a zscore > 3 on any of the top PCs (selected based on a cumulative variance cut off) or a z-score > 2 on its sample connectivity value calculated using the WGCNA47 package (v1.73).

### Identification of differentially expressed genes (DEGs)

For the identification of differentially expressed genes, we included samples from PPMI available whole-blood RNAseq data at month 6 across all groups (data release: 2021_v2_5release_0510). For *SNCA*+ versus PD, *GBA*+ versus PD, and *LRRK2*+ versus PD, differential expression analysis was performed using the same developed and consistent pipeline. We took as an input the AMP-PD derived feature count matrix for the pipeline. We initially applied a low-count filter to retain only genes with a count greater than zero out of the total 58,780 genes using the DEGfilter function from the DEGreport package (version 1.30.2)^13^. Subsequently, we employed the variancePartition package (version 1.30.2) to ascertain sources of variation^14^. Following the covariate correction pipeline, we constructed the DESeqDataSet object, adhering to package guidelines, and incorporating covariates such as ~ sex + neutrophils_mature + b_cells_naive + PCT_USABLE_BASES + genetic_status_enrollment. We performed two primary comparisons: (i) positive genetic cases (*SNCA*+, *GBA*+, and *LRRK2*+ cases) vs. sporadic PD, and (ii) unfavourable vs. favourable outcome in sporadic PD. For the identification of differentially expressed genes (DEGs), we used DESeq2 (version 1.40.2) with default settings^15^. Genes were considered significant if they achieved a False Discovery Rate (FDR)-adjusted *p*-value (*p*-adj) of less than 0.05. Up-regulated genes were identified when log_2_FoldChange exceeded 0.5 with a corresponding *p*-adj below 0.05, while down-regulated genes exhibited a log_2_FoldChange below 0.5 alongside a *p*-adj below 0.05.

### Functional Analysis of DEGs across comparisons

We carried out functional enrichment analysis using the Gene Ontology (GO) database (including Cellular Component (CC), Molecular Function (MF), and Biological Processes (BP), and Reactome databases. Functional enrichment analyses were performed and visualised using the R packages clusterProfiler (version 4.3.3)^16^ and ggplot2 (version 3.3.5)^17^, respectively. GO terms and Reactome pathways with a corrected p-value < 0.05 were considered significantly enriched by DEGs. For comparing biological functional profiles between study groups and time-points we used compareCluster function from the clusterProfiler package.

### Intersection analysis of DEGs with UpSetR

We used the UpSetR package (version 1.4.0) to visualize the overlap of significant DEGs across comparisons^18^. We combined into a single list object the DEG lists obtained for each comparison based on an adjusted *p*-value (<0.05) and log_2_FoldChange (0.5). We applied the upset function to compute and display all nonzero intersections among the sets, with the intersections ordered by descending frequency.

## RESULTS

### Description of demographic and clinical features across PD subtypes

We first explored key demographic and clinical features across *SNCA*+, *GBA*+, *LRRK2*+ and sporadic PD (sPD) cases summarized on **Table 1**. Clinically, the *SNCA*+ group comprised 26 cases (52.8% female) with the youngest mean age at diagnosis (47.2 ± 8.9 years), whereas *GBA*+ (n = 70) and *LRRK2*+ (n = 147) patients were diagnosed later (57.4 ± 9.6 and 58.9 ± 8.8 years, respectively), closely matching sPD (n = 387; 59.2 ± 9.2 years). Disease duration, calculated as age at last visit minus age at diagnosis, was longest in *SNCA*+ cases (8.09 ± 3.10 years) and shortest in *GBA*+ (5.80 ± 2.15 years), with *LRRK2*+ (7.08 ± 2.26 years) and sPD (7.70 ± 1.32 years) intermediate. Frequencies of dementia and mortality were highest in *SNCA*+ patients (30.8% for both outcomes), low in *GBA*+ (1.4% dementia; 14.3% mortality), and moderate in *LRRK2*+ (1.4% dementia; 5.4 % mortality) and sporadic PD (8.5% dementia; 10.6% mortality). Follow-up duration varied substantially: sporadic PD had the longest mean follow-up (87.9 ± 12.5 months), followed by *LRRK2*+ (52.3 ± 13.1 months), *SNCA*+ (46.8 ± 17.0 months), and *GBA*+ (29.3 ± 10.6 months).

**Table 1.** Description of demographics, clinical, follow-up and RNA-seq features in the Genetic Cohort and Sporadic PD cases from the PPMI study.

| Feature type | Features | Genetic Cohort - PD |  |  | Sporadic PD |
| --- | --- | --- | --- | --- | --- |
|  |  | *SNCA*+ | *GBA*+ | *LRRK2*+ |  |
| Demographics | Total cases, N | 26 | 70 | 147 | 387 |
|  | Sex (%), female | 14 (52.8) | 32 (45.7) | 77 (52.4) | 133 (34.4) |
|  | Age at baseline (mean, SD) | 51.5 (9.3) | 60.6 (8.9) | 61.7 (8.4) | 59.7 (9.3) |
| Clinical | Age at diagnosis (mean, SD) | 47.2 (8.9) | 57.4 (9.6) | 58.9 (8.8) | 59.2 (9.2) |
|  | Disease duration<sup>1</sup>, years (mean, SD) | 8.09 (3.1) | 5.8 (2.2) | 7.1 (2.3) | 7.7 (1.3) |
|  | Dementia cases (N, %) | 8 (30.8) | 1 (1.4) | 2 (1.4) | 33 (8.5) |
|  | Cases who died (N, %) | 8 (30.8) | 10 (14.3) | 8 (5.4) | 41 (10.6) |
| Follow-up | Follow-up<sup>2</sup>, months (mean, SD) | 46.8 (17.0) | 29.3 (10.6) | 52.3 (13.1) | 87.9 (12.5) |
|  | Month visit (max-min) | 0-61 | 0-49 | 0-73 | 0-109 |
| RNA-seq | Total samples, N (% availability) | 13 (50.0) | 25 (35.7) | 96 (65.3) | 259 (66.9) |
| <sup>1</sup> Disease duration is calculated as the difference between the age at last visit and age at diagnosis.<br><sup>2</sup> Follow-up duration is calculated as the time from the first visit month to the last visit month. |  |  |  |  |  |

In **Table 2**, we stratified each genetic and sporadic subgroup by disease outcome (favourable and unfavourable). We found that, across all PD subtypes, patients with an unfavourable outcome were older at baseline and at diagnosis than those with a favourable trajectory. We also noted that *SNCA*+ cases with unfavourable outcomes exhibited the longest disease duration, whereas *GBA*+ and *LRRK2*+ patients with poor prognosis had shorter disease courses. We did not observe differences in disease duration by prognosis in sPD. Finally, we observed that dementia prevalence was highest in unfavourable *SNCA*+ and sPD groups, followed by *LRRK2*+ and *GBA*+, and that mortality rates mirrored this pattern.

**Table 2.** Description of demographics features for favourable and favourable outcome cases per study arm in the PPMI cohort.

| Feature type | Features | Genetic Cohort - PD |  |  |  |  |  | Sporadic PD |  |
| --- | --- | --- | --- | --- | --- | --- | --- | --- | --- |
|  |  | SNCA+ |  | GBA+ |  | LRRK2+ |  |  |  |
|  |  | F | U | F | U | F | U | F | U |
| Demographics | Total cases, N | 12 | 14 (53%) | 48 | 22 (31%) | 107 | 39 (27%) | 264 | 122 (31%) |
|  | Sex (% female) | 7 (58.3%) | 7 (50.0%) | 22 (45.8%) | 10 (45.4%) | 48 (44.8%) | 28 (71.8%) | 87 (32.9%) | 46 (37.7%) |
|  | Age at baseline (mean, SD) | 49.9 (10.9) | 51.2 (10.1) | 60.0 (9.2) | 64.5 (8.4) | 61.1 (8.8) | 65.7 (8.1) | 58.7 (9.2) | 65.3 (8.8) |
| Clinical | Age at diagnosis (mean, SD) | 47.6 (11.0) | 46.8 (9.8) | 56.7 (9.7) | 61.5 (9.2) | 58.4 (9.2) | 62.3 (8.4) | 58.3 (9.1) | 64.7 (8.9) |
|  | Disease duration<sup>1</sup>, years (mean, SD) | 4.9 (4.0) | 8.1 (2.4) | 5.7 (2.5) | 5.6 (1.3) | 6.6 (2.4) | 5.6 (1.3) | 7.4 (1.3) | 7.4 (1.9) |
|  | Dementia cases (N, %) | - | 8 (57.1%) | - | 1 (4.5%) | - | 2 (5.1%) | - | 33 (22.0%) |
|  | Cases who died (N, %) | - | 8 (57.1%) | - | 10 (45.4%) | - | 8 (20.5%) | - | 41 (33.6%) |
| Follow-up | Follow-up<sup>2</sup>, months (mean, SD) | 31.8 (20.2) | 45.3 (17.0) | 24.7 (11.1) | 30.5 (13.7) | 45.7 (17.1) | 47.3 (15.5) | 84.7 (15.0) | 82.4 (19.4) |
|  | Month visit (max-min) | 0-61 | 0-61 | 0-49 | 0-49 | 0-61 | 0-61 | 0-109 | 0-109 |
| RNA-seq | Total samples, N | 4 | 9 | 15 | 10 | 70 | 25 | 179 | 80 |

Note: F (Favourable outcome) and U (Unfavourable outcome).
<sup>1</sup> Disease duration is calculated as the difference between the age at last visit and age at diagnosis.
<sup>2</sup> Follow-up duration is calculated as the time from the first visit month to the last visit month.

### Differential survival analysis across PD genetic subtypes

We performed a Kaplan-Meier survival analysis comparing *SNCA*+, *GBA*+, *LRRK2*+ and sporadic PD cases. In **Figure 1**, the y-axis represents the probability of remaining free from an unfavourable outcome (starting at 1.0) over disease duration in years (x-axis). A steeper decline in the survival curve indicates a higher likelihood of an unfavourable outcome. Notably, *GBA*+ patients exhibit the most pronounced decline in the survival curve, indicating the worst prognosis of the four groups. *SNCA*+ cases also show reduced survival, although the small sample size limits statistical power. Sporadic PD patients fare better than both *GBA*+ and *SNCA*+ groups but worse than *LRRK2*+ patients, whose curve declines most gradually. Consistent with these observations, hazard ratio (HR) values (**Table 3**) reveal that both *GBA*+ and SNCA+ cohorts carry an increased risk of unfavourable outcome relative to sporadic PD (HR > 1), reaching statistical significance for *GBA*+ group. In contrast, *LRRK2*+ patients have reduced hazard value (HR = 0.73), suggesting a lower risk compared to sporadic PD but not statistically significant.

**Figure 1.**
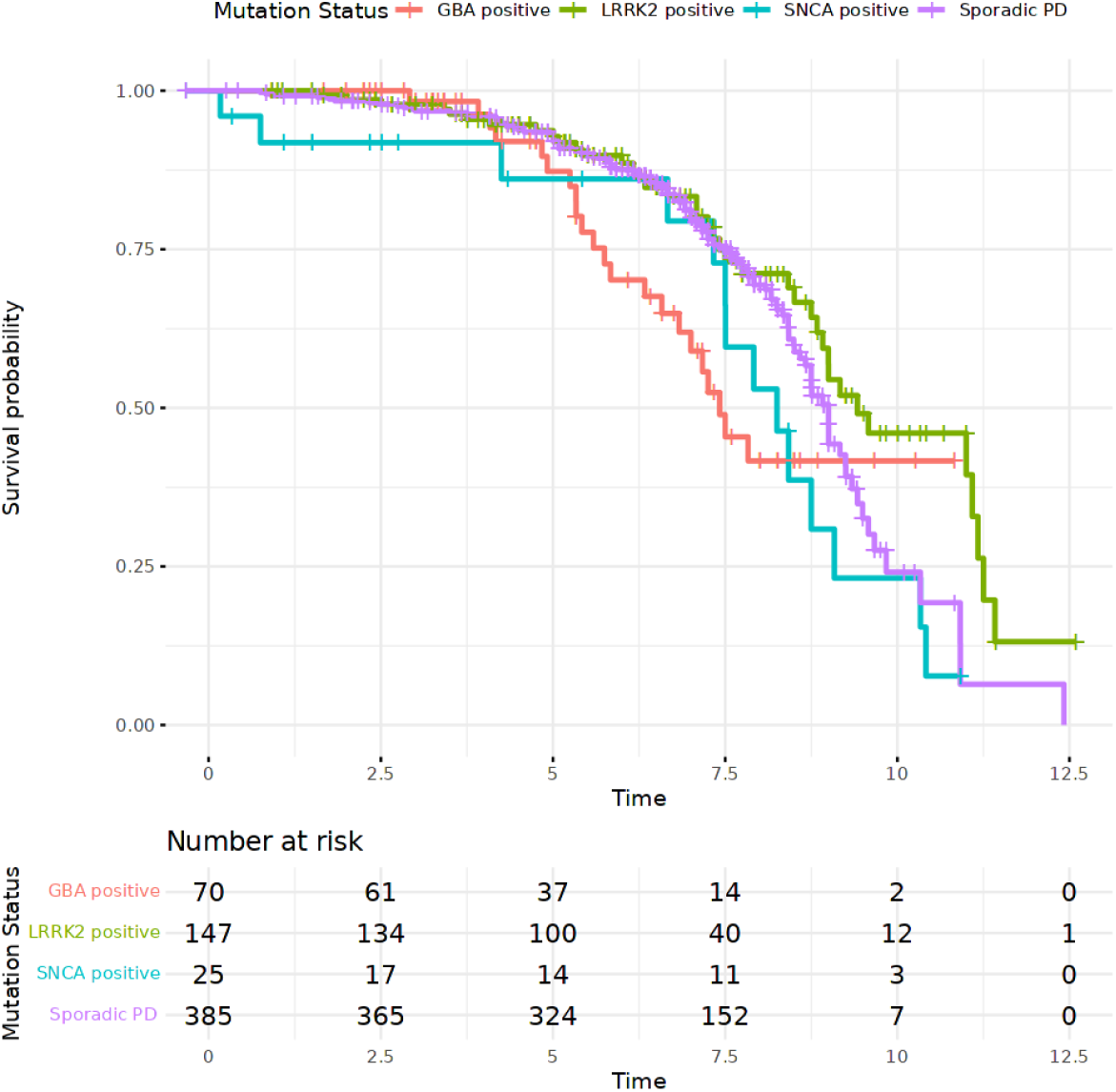
Kaplan–Meier survival curves showing time to clinical progression in Parkinson’s disease according to mutation status. Separate trajectories are shown for *GBA*-positive, *LRRK2*-positive, *SNCA*-positive, and sporadic cases. The y-axis represents survival probability, and the x-axis represents follow-up time (in years). The table below indicates the number of participants at risk at each time point for each group.

**Table 3.** Cox proportional hazards model results comparing SNCA+, GBA+ and LRRK2+ cases to the reference group (sporadic PD cases).

| **Genetic group** | **HR**<br>**(exp(coeff))** | **95% CI**<br>**(Lower-Upper)** | **p-value** | **Interpretation** |
| --- | --- | --- | --- | --- |
| ***SNCA+*** | 1.65 | (0.94-2.88) | 0.081 | Higher risk, but borderline significance. Small sample size. |
| ***GBA+*** | 1.61 | (1.02-2.53) | 0.042 | Higher risk vs Sporadic PD (statistically significant) |
| ***LRRK2+*** | 0.73 | (0.50-1.07) | 0.104 | Lower risk, but not statistically significant. |

### Differential gene expression analysis and functional enrichment analysis

We performed differential gene expression analysis at month 6 to define the specific signature related to genetically defined subgroups. We compared *SNCA*+, *GBA*+ and *LRRK2*+ cases against sPD (**Figure 2**; **Table 4**). We observed the fewest significant upregulated DEGs across positive mutation carriers in *SNCA*+ versus sPD (n=12), alongside a notable number of downregulated genes (n=98). We detected moderate transcriptomic changes in *GBA*+ against sPD, with 31 upregulated and 58 downregulated DEGs. The strongest impact emerged in *LRRK2* versus sporadic PD, where we identified 53 upregulated DEGs but a pronounced downregulation trend encompassing 250 genes. For the unfavourable and favourable outcome comparisons, we observed a small effect on gene expression with 6 upregulated DEGs and 2 downregulated DEGs (**Figure 2d**). Full lists of significant DEGs are provided in the Supplementary tables (**<u>Supplementary Table 1-4</u>**).

**Figure 2.**
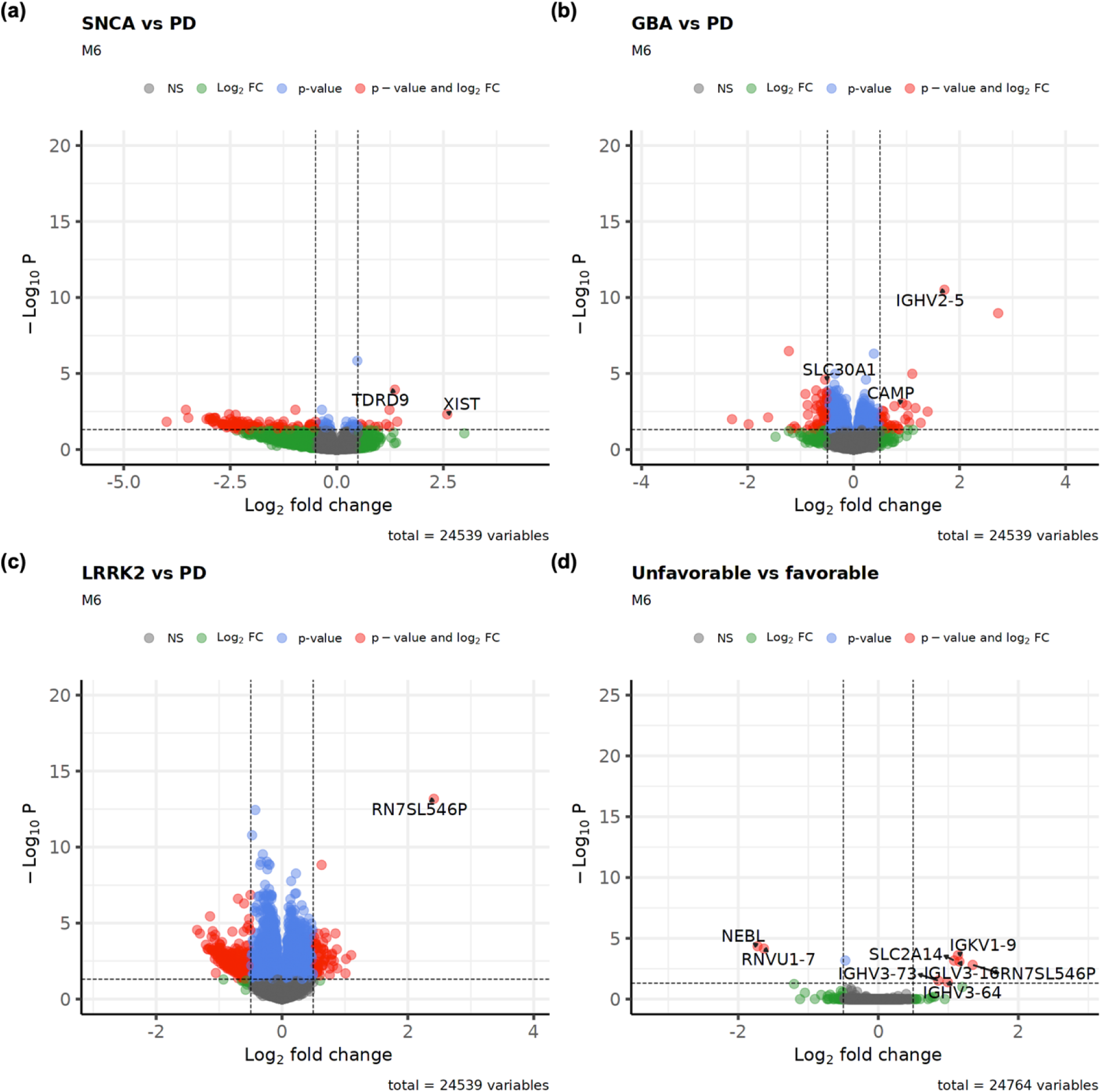
Volcano plots showing significant differentially expressed genes when comparing (a) *SNCA*, (b) *GBA* and (c) *LRRK2* positive carriers against sporadic PD cases. (d) DEGs when comparing unfavourable vs favourable sporadic PD cases. Plots show log transformed adjusted p-values (<0.05) on the y-axis against log2 fold change 0.5 on the x-axis.

**Table 4.** Distribution of upregulated and downregulated DEGs across comparisons.

| **Comparisons** | **Upregulated DEGs, N** | **Downregulated DEGs, N** | **Pattern interpretation** |
| --- | --- | --- | --- |
| ***SNCA+* vs PD** | 12 | 98 | The smallest number of upregulated genes across positive mutation carriers, but a substantial number of downregulated genes. |
| ***GBA+* vs PD** | 31 | 58 | More genes are downregulated than upregulated. |
| ***LRRK2+* vs PD** | 53 | 250 | The highest number of DEGs with a stronger downregulation trend suggesting a marked transcriptional impact. |
| **UP-PD vs FP-PD** | 6 | 2 | Small effect on gene expression. |

Figure 3 shows the top 20 significant enriched GO terms which describe molecular processes associated with the DEGs in each comparison. In **Figure 3a**, we mainly observed suppressed enriched GO terms related to cellular processes. In **Figures 3b and 3c**, we identified a high number of significant enriched terms related to the activation of the immune response in *GBA*+ and *LRRK2*+ when comparing with sPD. We observe a notable number of significant GO terms associated with the activation of the innate immune response in mucosa, antimicrobial humoral response or immunoglobulin production. We also analysed the overlap across significant GO enriched terms in all comparisons (**Supplementary Figure 1**). Interestingly, we observed an overlap of suppressed terms in *SNCA*+ vs sPD and *LRRK2*+ vs sPD and the highest overlap of activated terms between *GBA*+ vs sPD and *LRRK2*+ vs sPD.

**Figure 3.**
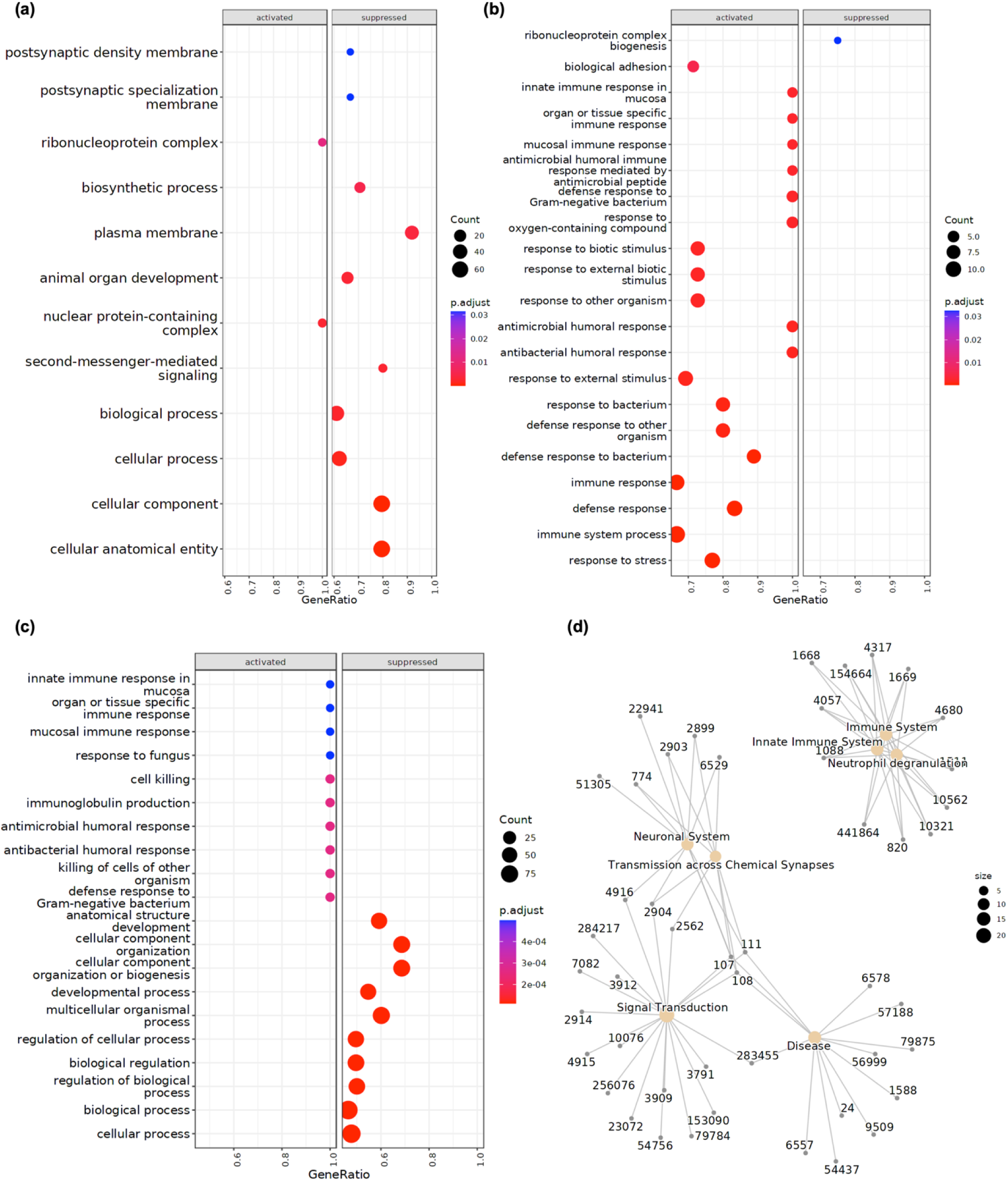
Functional enrichment dotplots showing significant Gene ontology enriched terms for (a) *SNCA*+ vs sPD, (b) *GBA*+ vs sPD and (c) *LRRK2*+ vs sPD. Network plot of Reactome enriched terms for LRRK2+ vs sPD. The size of each dot represents the GeneRation, the proportion of genes in the set associated with the given function. The dot color represents increasing statistical significance from darker blue to red (p-Adj). Figure 3. Functional enrichment dotplots showing significant Gene ontology enriched terms for (a) *SNCA*+ vs sPD, (b) *GBA*+ vs sPD and (c) *LRRK2*+ vs sPD. Network plot of Reactome enriched terms for *LRRK2*+ vs sPD. The size of each dot represents the GeneRation, the proportion of genes in the set associated with the given function. The dot color represents increasing statistical significance from darker blue to red (p-Adj).

### Intersection analysis between DEG sets

We performed an intersection analysis to explore the overlap of significant DEGs among PD subgroups. **Figure 4** shows an Upset plot summarizing the intersections of significant DEGs (p-adj < 0.05 and log_2_FoldChange = 0.10) from all comparisons. The largest overlap of upregulated DEGs (8 genes) appears to be shared between *GBA*+ vs PD and *LRRK2*+ vs PD (*CAMP, OLFM4, LINC02009, ABCA13, MMP8, CEACAM8, LTF, DEFA4*). Unique to *LRRK2*+ vs PD were 34 DEGs, while 15 DEGs for *GBA*+ vs PD and 10 DEGs for *SNCA*+ vs PD (**Figure 4a**). Among downregulated genes, *LRRK2* carriers showed the largest gene set (n=227), followed by *SNCA* (n=92) and *GBA* carriers (n=47) (**Figure 4b**). Notably, the largest overlap of downregulated DEGs (53 genes) is shared between *SNCA*+ vs PD and *LRRK2*+ vs PD, while more limited intersections were detected among genetic groups. Only a small subset of genes (*HYDIN, MYO18B*) was common to all comparisons, indicating that each group retains a partially distinct transcriptional signature. Full gene lists for each intersection are provided in **<u>Supplementary Table 5 and 6</u>**.

**Figure 4.**
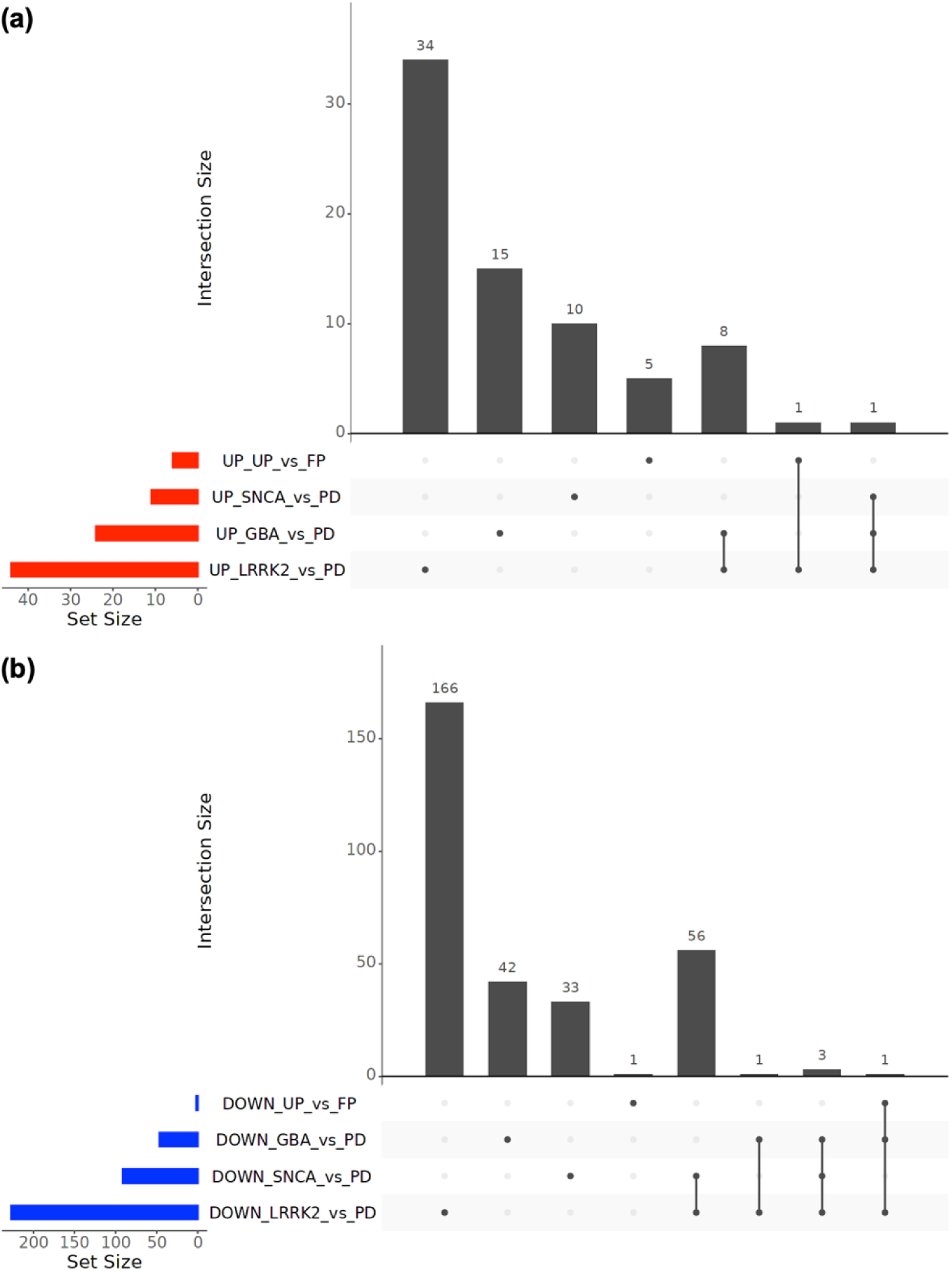
Visualization of the intersection between significant (a) upregulated and (b) downregulated DEG sets across *GBA*, *LRRK2* and *SNCA* mutation carriers compared to sporadic Parkinson’s disease patients.

## DISCUSSION

PD is increasingly recognized as a heterogeneous disorder with complex clinical manifestations and variable progression trajectories. Genetic factors such as mutations in *SNCA*, *GBA* and *LRRK2* contribute to distinct phenotypic subtypes, influencing age at onset, motor and non-motor symptomatology, and long-term outcomes. Understanding how these genetic differences shape disease courses is critical for developing better prognostic tools and personalized therapies.

We used data from well-characterized *SNCA*+, *GBA*+, *LRRK2*+, and sporadic PD patients under uniform inclusion criteria to minimize selection bias. Our Kaplan-Meier survival analysis confirmed longer survival in *LRRK2*+ and sporadic PD compared with *SNCA*+ and *GBA*+ carriers. These observations are in line with prior reports of earlier onset and more aggressive progression in *SNCA* mutation carriers, whereas *GBA* mutation carriers frequently present with non-motor symptoms such as dementia, compared to idiopathic PD patients^19^. However, the small *SNCA*+ sample size limits confidence in those comparisons and underscores the need for larger datasets in this subgroup. By contrast, *LRRK2*-associated PD, tends to have a slower motor progression but shows variable penetrance and phenotypic expressivity across carriers^20^.

In our differential expression analysis at month 6, we observed robust immune-related gene activation in both *LRRK2*+ and *GBA*+ patients even after adjusting for blood cell-type proportions. Notably, many downregulated DEGs overlapped between *LRRK2*+ and *SNCA*+ relative to sporadic PD, while a shared set of upregulated genes characterized the *LRRK2*+ and *GBA*+ comparisons. The blood-based changes in *GBA*+ and *LRRK2*+ early-phase patients point to systemic inflammatory processes that warrant further investigation. We did not detect significant transcriptional differences between unfavourable versus favourable outcome PD cases in blood, suggesting that differential gene expression may be more pronounced in affected brain regions than in peripheral samples.

Interestingly, the genes shared between *GBA* and *LRRK2* carriers include several transcripts previously linked to lysosomal function, mitochondrial metabolism, and vesicle trafficking, suggesting convergence on common cellular pathways despite different genetic origins. This pattern supports the notion that, while specific mutations shape distinct transcriptomic profiles, certain downstream processes may represent shared mechanisms of neurodegeneration across PD subtypes. Among the commonly downregulated genes, HYDIN and MYO18B were consistently decreased across all comparisons. Both genes are involved in structural and cytoskeletal processes rather than canonical dopaminergic pathways, suggesting that their regulation may reflect systemic or cell-structural changes rather than disease-specific neurodegenerative mechanisms.

By including estimated cell-type proportions in our models, specifically, “neutrophils_mature” and “b_cells_naive”, we demonstrated that the immune signature in *GBA*+ and *LRRK2*+ blood likely represents a true inflammatory activation. This supports previous studies implicating lysosomal dysfunction in *GBA* mutation carriers^21^ and innate immune dysregulation in *LRRK2* carriers in PD pathogenesis^22–24^. These findings support the potential utility of blood-based biomarkers for early detection and stratification of genetically defined PD subtypes.

In our study, we included deeply phenotypic genetic and sporadic PD cohorts with comprehensive covariate adjustment-including age at baseline, sex, disease duration and cell type proportions-thereby reducing confounding and improving the validity of our molecular insights. On the other hand, we acknowledge the limited number of *SNCA*+ samples, which constrains statistical power and interpretability for that subgroup. We plan to validate these transcriptomic signatures in larger, independent PD cohorts and to extend our analyses to postmortem brain samples. Functional studies dissecting the biological roles of the identified DEGs and pathways will be essential for translating our findings into novel therapeutic targets and precision-medicine approaches.

## CONCLUSION

In conclusion, our integrated clinical and transcriptomic analysis of *SNCA*, *GBA* and *LRRK2*-mutation carriers and sporadic PD cases highlight both shared and mutation-specific features of disease progression. We demonstrate that *SNCA*+ patients exhibit earlier onset and poorer survival, while *LRRK2*+ and sporadic PD cohorts follow more benign trajectories. Blood-based transcriptomic profiling reveals prominent immune activation in *GBA*+ and *LRRK2*+ carriers suggesting systemic inflammation as an early hallmark of these genetic subtypes. Although peripheral expression changes did not distinguish unfavourable from favourable outcome patients, our findings underscore the value of combining survival modelling with molecular signatures to refine PD stratification. Moving forward, larger cohorts and validation in brain tissue will be critical to identify molecular mechanisms underlying distinct PD subpopulations.

## Data Availability

All data produced in the present study are available upon reasonable request to the author.

## ACKNOWLEDGMENTS

Data used in the preparation of this article were obtained on [2023-09-03] from the Parkinson’s Progression Markers Initiative (PPMI) database (www.ppmi-info.org/access-data-specimens/download-data), RRID:SCR_006431. For up-to-date information on the study, visit www.ppmi-info.org. For up-to-date information on the study, visit www.ppmi-info.org. PPMI – a public-private partnership – is funded by the Michael J. Fox Foundation for Parkinson’s Research and funding partners, including AbbVie, Allergan, Amathus Therapeutics, Avid Radiopharmaceuticals, Biogen, BioLegend, Bristol Myers Squibb, Celgene, Denali, GE Healthcare, Genentech, GlaxoSmithKline, Golub Capital, Handl Therapeutics, Insitro, Janssen Neuroscience, Lilly, Lundbeck, Merck, Meso Scale Discovery, Neurocrine Biosciences, Pfizer, Piramal, Prevail Therapeutics, Roche, Sanofi Genzyme, Servier, Takeda, Teva, UCB, Verily and Voyager Therapeutics. Funding: ALGM MJFF grant, RR, HM Team Hardy and GP2 funding. Team Hardy - This research was funded in part by Aligning Science Across Parkisnons [ASAP 000478] through Michael J. Fox Foundation for Parkinsons Research (MJFF).

## Supplementary Material

### Supplementary tables

**Supplementary Table 1 List of Differentially Expressed Genes SNCA+ vs sPD at M6.**

**Supplementary Table 2 List of Differentially Expressed Genes GBA+ vs sPD at M6.**

**Supplementary Table 3 List of Differentially Expressed Genes LRRK2+ vs sPD at M6.**

**Supplementary Table 4 List of Differentially Expressed Genes unfavourable vs favourable outcome of PD cases at M6.**

**Supplementary Table 5. Overlapping upregulated genes across *GBA*, *LRRK2*, and *SNCA* mutation carriers compared to sporadic Parkinson’s disease.**

**Supplementary Table 6. Overlapping downregulated genes across *GBA*, *LRRK2*, and *SNCA* mutation carriers compared to sporadic Parkinson’s disease.**

### Supplementary figures

**Supplementary Figure 1.**
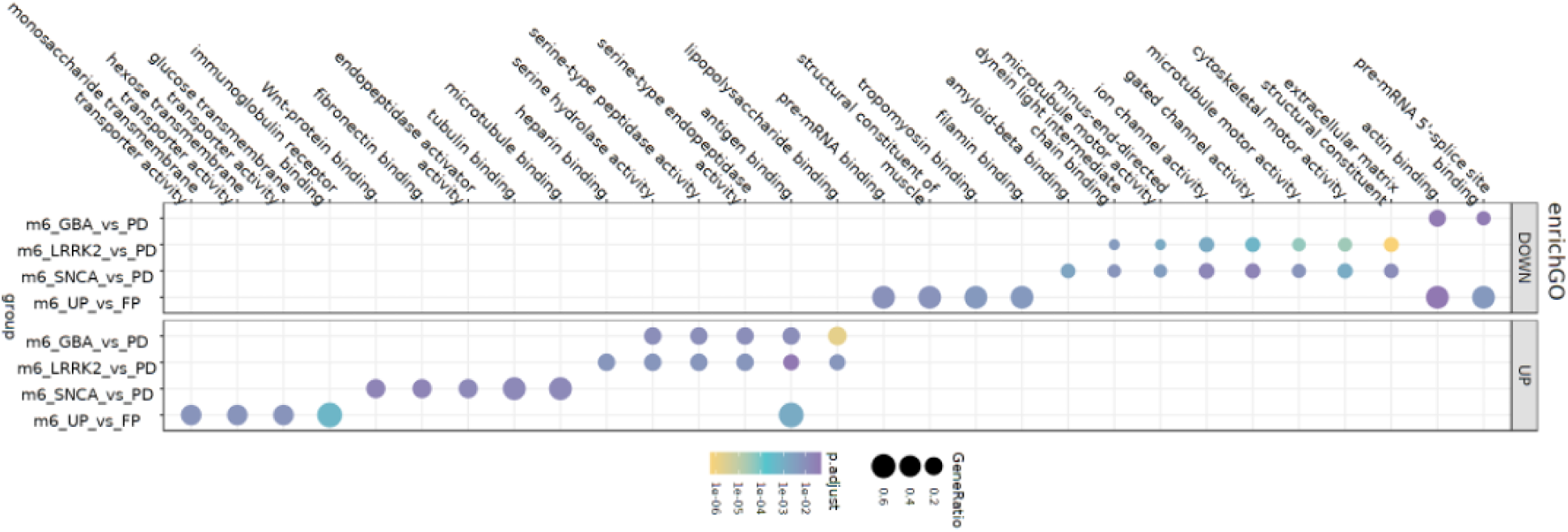
Dotplot comparing clusters of significant GO enriched terms when comparing *SNCA*+, *GBA*+ and *LRRK2* vs sPD and unfavourable and favourable PD cases. The size of each dot represents the GeneRation and the dot color represents increasing statistically significant from purple to yellow (p-Adj < 0.05).

